# Precision synbiotics increase gut microbiome diversity and improve gastrointestinal symptoms in a pilot open-label study for autism spectrum disorder

**DOI:** 10.1101/2022.10.26.22281525

**Authors:** Joann Phan, Diana C. Calvo, Divya Nair, Suneer Jain, Thibaut Montagne, James Corbitt, Kelsey Blanchard, Shirin Treadwell, James Adams, Rosa Krajmalnik-Brown

## Abstract

The use of prebiotics and probiotics to improve symptoms associated with autism spectrum disorder (ASD) has varied from study to study, indicating the complex and heterogeneous nature of the disorder and the behaviors and gastrointestinal symptoms associated with ASD. There is a wide variety in the severity of symptoms and developmental impediments across the population. Gut microbiome studies have also shown unique but varied microbial signatures in ASD. While there have been successes in pre-clinical and clinical trials with prebiotic and probiotic components, the limited population sizes have promising yet inconclusive results. This study addresses this issue by 1) enrolling an ASD cohort of 296 children and adults and comparing their deep DNA metagenomic sequencing of gut microbiomes to that of an age-matched neurotypical cohort and 2) individually formulating a precision synbiotic (probiotic and prebiotic) tailored towards each individual’s needs and conducting pre/post evaluations of ASD and GI symptoms and longitudinal whole genome microbiome sequencing. At baseline, there was significantly lower microbiome diversity in the ASD group relative to controls. Microbes, pathways, and gene families significantly differed between the two populations. The ASD microbiome had higher abundances of pathogens, such as *Shigella, Klebsiella, Mycobacterium*, and *Clostridium*, but lower abundances of beneficial microbes, including *Faecalibacterium*. With a 3-month synbiotic supplementation, the microbiome diversity of the 170 ASD participants completing the study increased and became closer to the neurotypical controls. Significant shifts in microbial and pathway abundances were also measured at the second ASD timepoint. In addition to changes in the gut microbiome, there was a significant reduction in gastrointestinal discomfort. There were also improvements in some ASD-related symptoms; however, we cannot exclude that these were potentially due to the open-label nature of the study. Changes in the gut microbiome composition and functional capacity, along with a reduction in gastrointestinal symptoms and potential changes in behavior, highlight the importance of metagenomics, longitudinal studies, and the potential for therapeutic microbial supplementation in ASD.

## Introduction

Autism spectrum disorder (ASD) is a complex neurodevelopmental condition characterized by significant impairments in social interaction and communication, repetitive and restricted behaviors, and deficits in sensory reactivity (Khaleghi et al., 2020). Although there are wide variations in the severity of symptoms among ASD patients, gastrointestinal symptoms are a common comorbidity. The percentage of patients with ASD presenting gastrointestinal (GI) issues varies from study to study. Still, there is a general trend that ASD patients often suffer from diarrhea, constipation, bloating, gastroesophageal reflux, and/or abdominal pain (Manokaran and Gulati 2022; D.-W. Kang et al. 2020; Adams, Johansen, et al. 2011). The high frequency of GI problems in ASD patients raised a question about the relationship between autism-related symptoms and the gut microbiome (Alharthi et al., 2022). Many studies have accumulated evidence that dysbiosis might affect behavioral and gastrointestinal symptoms in children with ASD (Fattorusso et al. 2019; Sharon et al. 2019; Principi and Esposito 2016; Krajmalnik-Brown et al. 2015a; Qureshi et al. 2020). The bidirectional relationship between the gut and the brain, known as the gut-brain axis, is a widely accepted concept (Suganya and Koo 2020).

Although most studies conclude that patients with ASD have a distinctive gut microbiome, there is inconsistency in the taxa reported between these studies. Some studies show no differences between patients with ASD and neurotypical cohorts. This inconsistency is partially explained by the complexity of ASD, given the heterogeneity of symptoms and their severity (Fouquier, Moreno Huizar, et al. 2021). However, there are several more reasons that may explain the differences seemed between studies: 1) the methodological approach to collect data is drastically different between studies; 2) the diagnosis of GI symptoms is not standardized; 3) small cohorts in most studies make reproducibility challenging; 4) differences in the diet; 5) differences in geographical area/country, including different bacteria in those regions; and 6) differences in race/ethnicity (Zhang et al. 2022; Fouquier, Moreno Huizar, et al. 2021; Krajmalnik-Brown et al. 2015b). A non-invasive study analyzed the diet, fecal microbiome, and fecal consistency of 247 children, where 99 children were diagnosed with ASD (Yap et al. 2021). The authors concluded that there are negligible associations between the gut microbiome and ASD diagnosis and that the gut microbiome seen in patients might be associated with dietary preferences. However, the authors linked ASD restrictive interest with a less diverse diet and lower beta and alpha diversity in fecal metagenomics.

The gut-brain axis concept has triggered attempts to modulate the gut microbiome with different approaches, such as fecal microbiota transplantation (FMT), probiotics, prebiotics, and synbiotics (a combination of probiotics and prebiotics (Tan et al. 2021). An open-label study of microbiota transfer therapy (an intensive version of FMT) reported substantial improvement in behavioral and gastrointestinal symptoms in 18 children with ASD even after two years after treatment and also found an increase in *Bifidobacterium, Prevotella*, and *Desulfovibrio* of the gut microbiome (D. W. Kang et al. 2017; 2019). The results from trials with pre/probiotics have differed from study to study, some with inconclusive findings mainly due to small cohorts. For example, (H. M. R. T. Parracho et al. 2010) showed an increase of *Enterococci* and *Lactobacilli* and a decrease of *Clostridium* when administered a *Lactobacillus* strain to children with ASD. However, the authors acknowledge the very high dropout of individuals and the statistical weakness of the study. In a cohort of 10 children with ASD, (Tomova et al. 2015)) also found changes in the gut microbiome of children with ASD after four months of administration of a probiotic that included three strains (*Lactobacillus, Bifidobacterium* and *Streptococcus*) thrice a day showed a decrease of *Bacteroidetes* and *Firmicutes* after treatment but an increase in the *Bacteroidetes*/*Firmicutes* ratio. Shaaban et al. 2018 presented a cohort (n=30) with different results; they found an increase in *Bifidobacterium* and *Lactobacilli* after a 3-months treatment of probiotics containing these strains (Shaaban et al. 2018).

Some studies focused on the effects on behavior and GI symptoms while using probiotics, with inconsistent findings. (Kałuzna-Czapli^ń^ska and Błaszczyk 2012) reported no changes in GI symptoms after a 2-month treatment with *Lactobacillus* administered twice daily in 22 children with ASD; however, they found lower D-arabinitol levels in their urine after treatment. D-arabinotol is a sugar alcohol that is measured to detect candidiasis. Grossi et al. identified a decrease in abdominal symptoms after administering a 9-strain probiotic to a 12-year-old child for three and ten months after treatment (Grossi et al. 2016a). Eugene et al. also improved GI symptoms after administering VISBIOME to 13 children with ASD who retained *Lactobacillus* (Eugene Arnold et al. 2019). Similarly, Santocchi et al. found improvements in GI symptoms and improvements in adaptative functioning and sensory profiles when 42 children with ASD were administered Vivomixx® probiotics and 43 children with a placebo for six months (Santocchi et al. 2020). Liu et al. also found improvements in behavior and anxiety after administering *Lactobacillus plantarum* PS128 to 35 children with ASD with a placebo group of 36 children (J. Liu et al. 2019). A similar size group was assessed, where 37 children were treated with a 6-strain probiotic for four weeks and found a decrease in the Autism Treatment Evaluation Checklist score [ATEC] (Niu et al. 2019).

Two studies used prebiotics exclusively. A cohort of 13 children with ASD showed improvement in GI symptoms after ingesting hydrolyzed guar gum for 2-15 months (Inoue et al. 2019). Grimaldi et al. had a slightly different approach, analyzing the effects of restricted and un-restricted diets combined with a 6-week Bimuno® galactooligosaccharide treatment in 30 children with ASD (Grimaldi et al. 2018). They found that a restrictive diet combined with the prebiotic showed a significant improvement in social responsiveness (Grimaldi et al. 2018).

Finally, two other studies used synbiotics (a combination of probiotics and prebiotics). One study administered four probiotic strains (two *Lactobacillus* and two *Bifidobacterium*) with fructooligosaccharide to 16 children with ASD, finding significant improvements in behavioral and gastrointestinal symptoms (Y. Wang et al. 2020). Similarly, the other study combined *Bifidobacterium infantis* with bovine colostrum product and treated eight children with ASD. They were randomly assigned to receive either the combination or the prebiotic exclusively (Sanctuary et al. 2019). Despite some observations of slight improvement in GI symptoms in the treated group, the sample size was too small to be conclusive. In summary, there have been many studies of a variety of pre/probiotics, with most involving small open-label studies of small cohorts. Some of the studies found improvement in GI and ASD symptoms.

However, few studies have evaluated the effect of pre/probiotics on the microbiome. Since there are wide variations in the microbiome of individuals with ASD, we believe synbiotic supplementation should be customized for the individual. One company, Sun Genomics, develops customized probiotics for individuals based on the results of metagenomic analysis from fecal samples and a personal health and diet survey of each person (https://Flore.com/). Their product, Floré^®^, is a personalized combination of typically 1-2 prebiotics and 4-8 probiotics chosen from a list of more than 100 possible ingredients to balance the individual’s gut microbiome. The objective of this work was to investigate the baseline characteristics of a large cohort of children and adults with ASD vs. healthy controls and to investigate the effect of individually customized probiotics on the GI and ASD symptoms of individuals with ASD. Here, we present a novel approach to pre/probiotic treatment for children with ASD by administering a synbiotic formula customized for each person from a repository of more than 100 ingredients. The gut microbiome was evaluated at baseline by deep whole-genome metagenomic sequencing and at approximately three months after treatment, including taxonomy, pathways, and genes. Data on health history, autism-related symptoms, social responsiveness, anxiety, GI symptoms, diet, and allergies, were collected before and after treatment. Fecal samples from 123 neurotypical children were also evaluated with whole-genome sequencing for taxonomy, pathways, and gene family classification for comparison with the ASD cohort.

## Materials and Methods

### Participants and sample collection

Participants were invited to participate in this study if they were new customers of Sun Genomics and had purchased Floré^®^ but had not yet started treatment. They viewed a study ad and then a written consent form that explained the study and their potential role and invited them to participate. The consent form was signed by the parent of the person with ASD or by the adult with ASD if they did not have a legal guardian. The study was approved by the IRB of Arizona State University as STUDY00012299 and registered on clinicaltrials.gov prior to enrolling participants.

The inclusion criteria for the ASD group included the following:

1) A new client of Sun Genomics (has applied for testing and treatment but has not yet begun treatment)

2) Diagnosis of ASD (documentation of a clinical diagnosis of ASD) was obtained from participants and then verified by an evaluation of the Social Responsiveness Scale (SRS-2) with a raw score above 60.

3) Children and adults ages 2.5-75 years

The exclusion criteria included:

1)Antibiotic use in the last two months (not counting topical antibiotics)

2) Any changes in medications, nutritional supplements, or therapies in the last two months or any plans to change them during the first three months of probiotic treatment.

A total of 296 participants with ASD were included in this study (Table 1). The ASD group was matched with a group of 123 healthy neurotypical, age-matched controls from the Sun Genomics customer base to compare their whole-genome sequencing of their fecal sample. Based on their health and diet survey, the controls were further screened for having no neurodisabilities, serious gastrointestinal disorders, or disease.

**Table 1.**
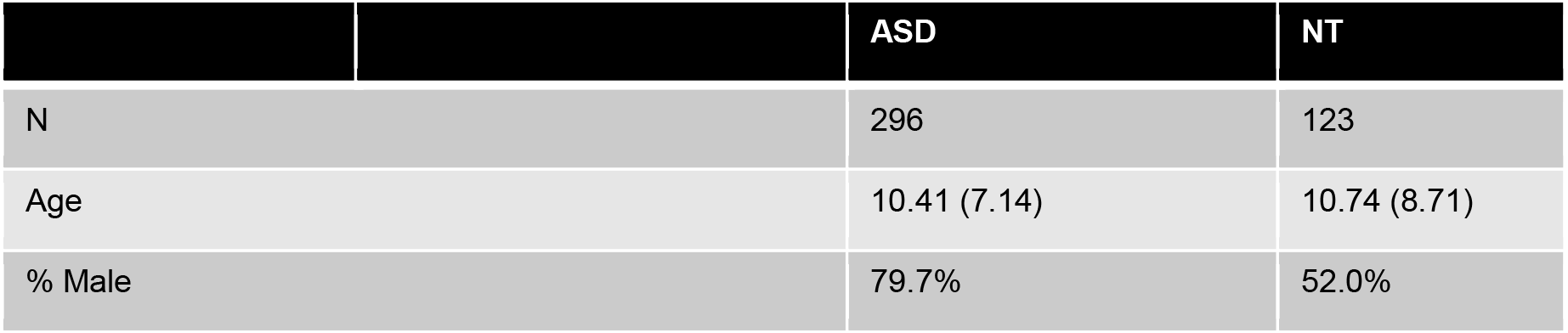
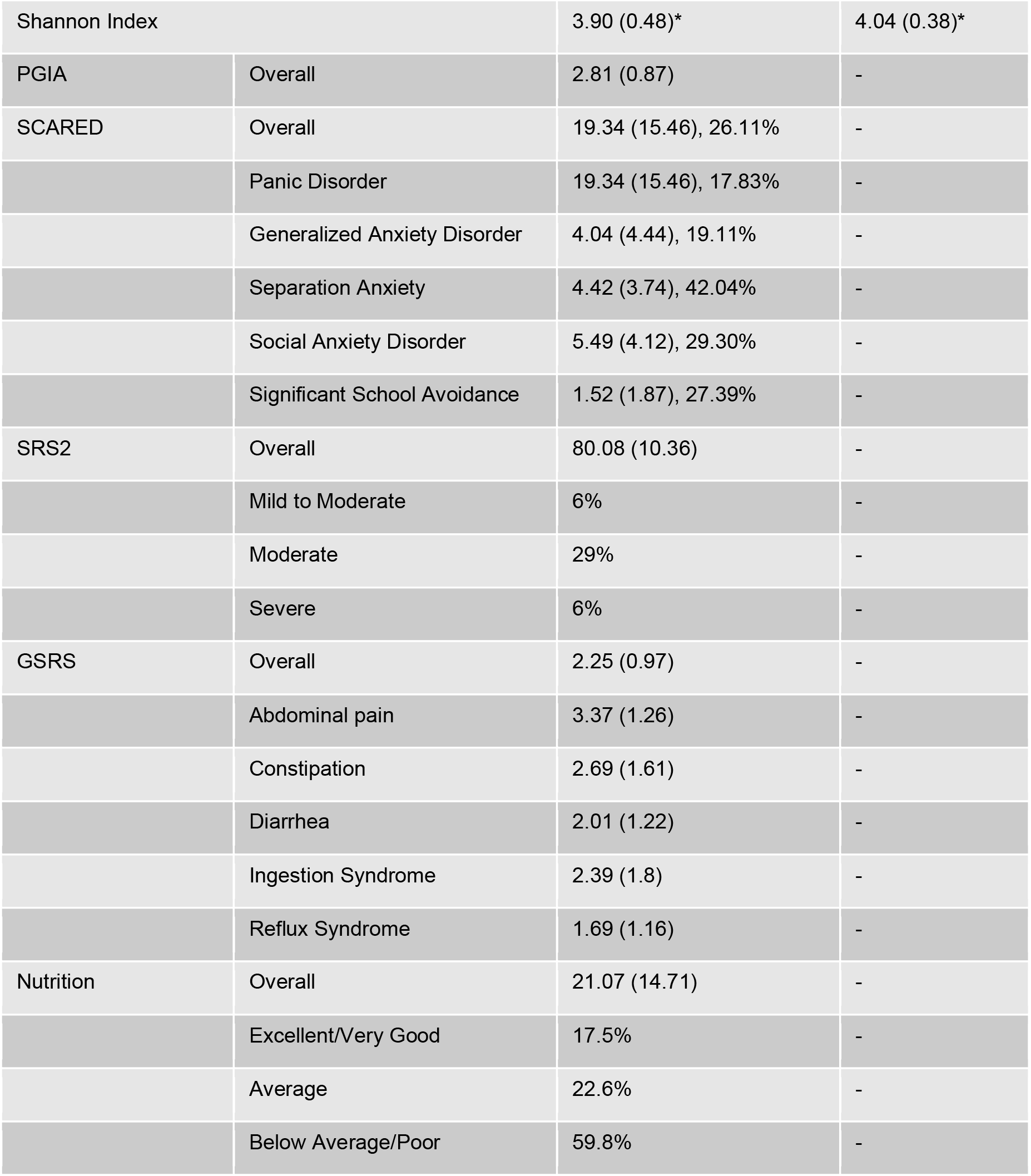
Study cohort demographics. Listed are the number of subjects, age, and gender of each cohort and the summaries of surveys taken by ASD study participants. The PGIA survey is the parental global impressions of autism. SCARED is the screen for childhood related anxiety disorders. SRS2 is the social responsiveness scale. GSRS is the gastrointestinal symptom response scale. The nutrition survey assesses dietary habits and the number of daily servings of various food categories. Values within the parentheses indicate SD from the mean. Percentages indicate the population proportion that falls within a subcategory (SCARED) or indicate a specific condition or phenotype (SRS2 and nutritional assessment surveys).

|  | ASD | NT |
| --- | --- | --- |
| N | 296 | 123 |
| Age | 10.41 (7.14) | 10.74 (8.71) |
| % Male | 79.7% | 52.0% |

|  |  |  |  |
| --- | --- | --- | --- |
| Shannon Index |  | 3.90 (0.48)* | 4.04 (0.38)* |
| PGIA | Overall | 2.81 (0.87) | - |
| SCARED | Overall | 19.34 (15.46), 26.11% | - |
|  | Panic Disorder | 19.34 (15.46), 17.83% | - |
|  | Generalized Anxiety Disorder | 4.04 (4.44), 19.11% | - |
|  | Separation Anxiety | 4.42 (3.74), 42.04% | - |
|  | Social Anxiety Disorder | 5.49 (4.12), 29.30% | - |
|  | Significant School Avoidance | 1.52 (1.87), 27.39% | - |
| SRS2 | Overall | 80.08 (10.36) | - |
|  | Mild to Moderate | 6% | - |
|  | Moderate | 29% | - |
|  | Severe | 6% | - |
| GSRS | Overall | 2.25 (0.97) | - |
|  | Abdominal pain | 3.37 (1.26) | - |
|  | Constipation | 2.69 (1.61) | - |
|  | Diarrhea | 2.01 (1.22) | - |
|  | Ingestion Syndrome | 2.39 (1.8) | - |
|  | Reflux Syndrome | 1.69 (1.16) | - |
| Nutrition | Overall | 21.07 (14.71) | - |
|  | Excellent/Very Good | 17.5% | - |
|  | Average | 22.6% | - |
|  | Below Average/Poor | 59.8% | - |

The ASD group completed extensive baseline evaluations regarding dietary and nutritional habits, behavior, birth and infancy history, allergies, social responsiveness, anxiety-related disorders, and gastrointestinal symptoms. The surveys collected were 1) Parent Global Impressions of Autism (PGIA), 2) Screen for Childhood-Related Anxiety Disorders (SCARED), 3) Social Responsiveness Scale (SRS-2), 4) Gastrointestinal Symptom Rating Scale (GSRS), and 5) nutritional assessment. The PGIA questionnaire assesses observable behaviors in children with ASD, including language, cognition, play skills, sociability, hyperactivity, and others. On average, the cohort had moderate severity of symptoms (Table 1). The SCARED survey assesses five different classes of anxiety, including panic disorder, generalized anxiety disorder, separation anxiety, social anxiety disorder, and significant school avoidance. Each of the categories has a scoring threshold that may suggest the presence of an anxiety disorder.

The nutritional assay asks for the number of servings of different food types consumed daily and common dietary habits. Each question has a positive or negative value depending on the kind of food assessed. For example, servings of nuts and seeds have a positive value, while refined grains have a negative score based on daily consumption. Other questions consider whether the daily diet includes artificial colors, flavors, or additives and whether the diet is organic. Diet quality was determined by summing up the score of the survey. Higher scores are associated with excellent or good diet quality, and lower scores are associated with poorer diets.

Fecal samples were collected via the Floré^®^ research edition kit for metagenomic sequencing. The Fecal sample was organized by the parent or participant of the study with provided kit instructions. Fecal samples from the control population were collected with the gut microbiome test kit (Floré^®^ gut health test kit). Samples are collected in a proprietary stabilization buffer and on a swab and shipped 2-day at ambient temperature.

As part of the longitudinal study with precision synbiotic supplementation, an additional timepoint from the ASD population was collected after three months of the supplementation. Different for each participant, formulations included 4 to 8 probiotic strains and 1 to 2 prebiotics, each at different concentrations. Participants were instructed to take it daily. On average, there was a 5 to 6-month difference between the generation of the first baseline timepoint and the second timepoint sample receipt and processing. Of the ASD cohort, there was a total of 170 participants with a second timepoint. GSRS, PGIA, SCARED, and SRS2 follow-up surveys were collected again upon the second fecal sample collection.

### Metagenomic sequencing and bioinformatic analysis

DNA extraction and library preparation methods are previously described in Phan et al. 2021. Briefly, DNA was extracted and purified with a proprietary process (patents 10428370 and 10837046). Library prep and size selection was performed with NEBNext^®^ reagents and MagJet^®^ magnetic beads, respectively. Library quantitation was performed with qPCR prior to normalization. Libraries were sequenced on an Illumina^®^ NextSeq 550 (Illumina^®^, San Diego, CA). Once sequenced, reads were quality filtered and processed to remove human reads. For taxonomic analysis, reads were aligned to a hand-curated database of 23,000 microbial species. Metabolic pathways and gene families were determined using HUMAnN2 with the MetaCyc and UniRef90 databases.

### Statistical analyses

The downstream statistical analyses in this paper were performed in R. Permutational multivariate analysis of variance (PERMANOVA) with the “adonis” function from the “vegan” package. Bray-Curtis dissimilarity was calculated with the “vegan” package and was used for principal-coordinates analysis. Pearson correlations were used to investigate parametric relationships between variables of interest. Random forest analysis was performed to identify variables of importance between the two cohorts in the microbiome, pathways, and gene family datasets. Subsequent significance testing with FDR corrections was performed on the random forest variables of importance to compare the relative abundances of microbes from each cohort. The Mann-Whitney U test or Wilcoxon rank sum tests were performed to determine statistical significance between observations. FDR corrections were used to control for multiple hypothesis comparisons. The microbial abundance data were rarefied, and the sample sum was normalized to account for differences in sequencing depth. The pathway and gene family datasets were sample sum normalized to account for differences in sequencing depth.

## Results

### Subject demographics and behavioral, gastrointestinal, and nutritional survey summaries

A total of 296 study participants with ASD and 123 neurotypical subjects were included in this study (Table 1). All subjects in the ASD cohort obtained and submitted clinical diagnosis documents for participation in the study. ASD and control cohorts were age-matched, with mean ages of 10.41 and 10.74, respectively. The percent male in the ASD cohort was 79.7%, while the control was 52.0% (Table 1). An initial analysis found no significant differences in microbial alpha or beta diversity between sex in either cohort, so the control group was not restricted to match the gender of the ASD group.

Using the information collected from the surveys, 26% of the ASD population may have an anxiety disorder, as assessed by the SCARED survey (Table 1). As for each specific anxiety disorder, 17% to 42% of the study participants may have the presence of at least one of the anxiety disorders (Table 1). The SRS2 survey indicates that 64% of the ASD cohort has severe difficulties in social responsiveness (Table 1). A smaller proportion of the cohort has mild to moderate and moderate difficulties, with 6% and 29%, respectively (Table 1).

From a rating scale of no discomfort to very severe discomfort, the ASD cohort, on average, had slight to mild discomfort as assessed with the GSRS survey (Table 1). Abdominal pain had a higher average subtype score with mild to moderate discomfort, while reflux syndrome was lower with no to slight discomfort (Table 1). The nutritional survey found that approximately 60% of the ASD cohort had below average or poor diets, 22.6% had average, and 17.5% had excellent or very good diets (Table 1). Higher scores calculated from the nutritional survey are associated with good diets and lower scores are associated with poor diets (also explained in Methods).

### Microbial diversity differed in the ASD cohort relative to controls and improved after probiotic treatment

To compare microbial diversity between the ASD and control cohorts, alpha and beta diversity metrics were calculated. The ASD cohort had a significantly lower Shannon score and microbial evenness than the control cohort (Figure 1a). However, the difference was small compared to the wide variation within each group. There was no significant difference in beta diversity as calculated by a PERMANOVA based on a Bray-Curtis dissimilarity matrix (p value 0.065 and R^2^ = 0.00319). The microbiome beta diversity was also displayed with principal coordinates analysis. There were significant differences in the PCO1 and PCO2 coordinates of the ASD and NT baseline cohorts (Wilcoxon test, p value = 0.0102 and 4.68e-12, respectively, Figure 1).

**Figure 1.**
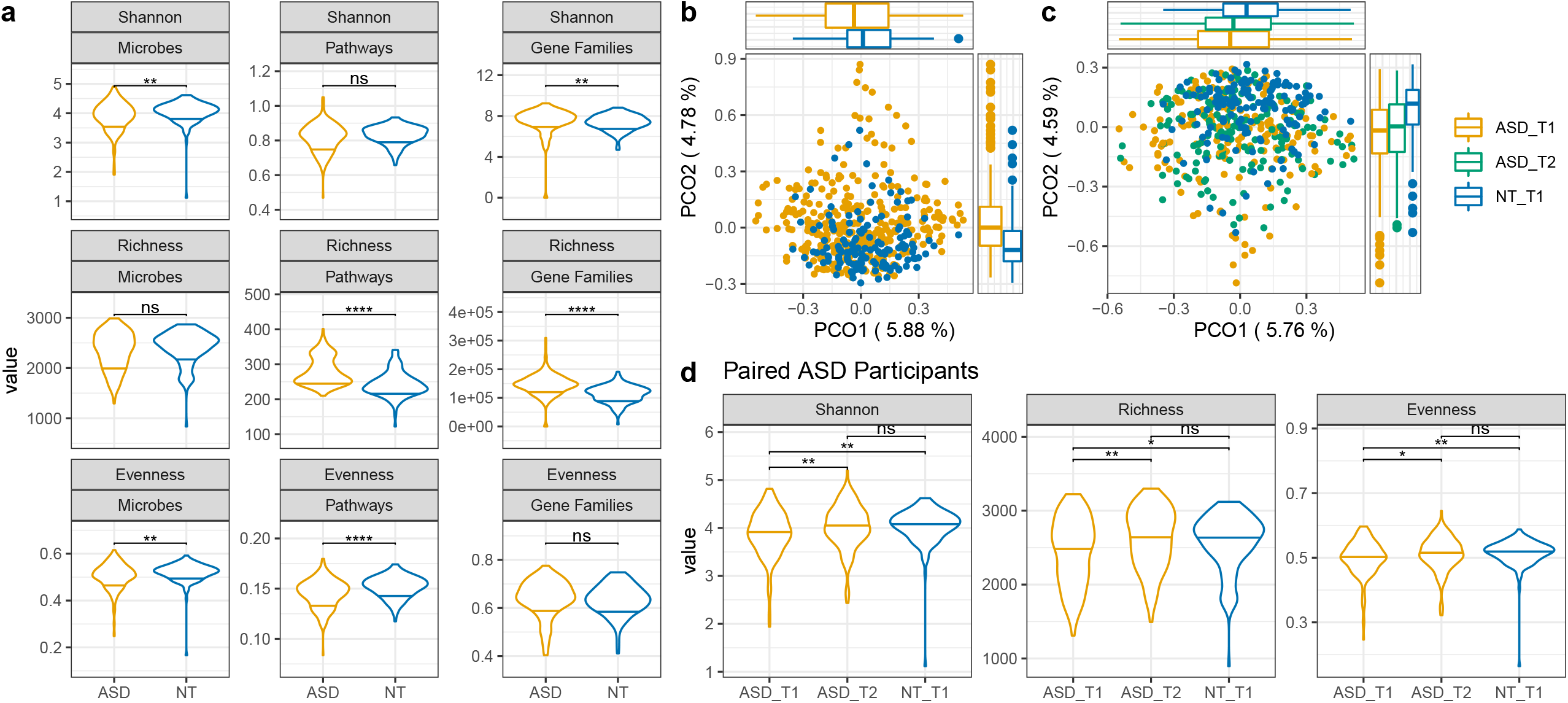
Diversity comparisons between ASD and neurotypical cohorts and paired longitudinal ASD subjects. A) Shannon index, richness, and evenness across microbiome composition, metagenomic pathways, and gene families between the two cohorts. B) PCO of the ASD and neurotypical cohorts. Yellow dots represent ASD, and blue dots represent NT. The boxplots along each axis represent the distribution points within the PCO. The Wilcoxon test for comparisons between cohorts resulted in a p-value = 0.0102 for PCO1 and p-value = 4.68e-12 for PCO2. C) PCO of paired ASD timepoint 1 and 2 and neurotypical timepoint one samples. For PCO1, ASD_T1 vs NT_T1 p.adj = 0.006, ASD_T2 vs NT_T1 p.adj = 0.0165. For PCO2, ASD_T1 vs NT_T1 p.adj = 2.43e-9, ASD_T1 vs NT_T1 p.adj = 2.62e-8. Principal coordinates were calculated based on a Bray-Curtis dissimilarity matrix. D) Alpha diversity at baseline (timepoint 1) for ASD and neurotypical and the subsequent timepoint 2 for the ASD cohort. Wilcoxon rank sum tests were performed for all tests with Benjamini-Hochberg post-hoc corrections for multiple comparisons. * p-value < 0.05; ** p-value < 0.01; **** p-value < 0.0001; ns not significant.

The analysis included 170 ASD participants with data for baseline and after three months of supplementation with Floré^®^. In the paired samples, there was a significant increase in the three assessed alpha diversity indexes (Shannon, richness, and evenness) in timepoint two relative to timepoint one at baseline (Figure 1), and the microbial alpha diversity of ASD timepoint 2 was not significantly different compared to the neurotypical cohort (Figure 1d). There was a significant difference between the PCO1 and PCO2 axes between both timepoints of ASD and the NT baseline (Wilcoxon test, p.adj < 0.05, Figure 1). There was no significant difference between ASD timepoints 1 and 2 (Figure 1b and 1c).

### Species evenness and richness correlated negatively with anxiety and positively with daily servings of fruit

To investigate whether differences in microbial alpha diversity in the ASD cohort were related to any of the observed behaviors, nutrition, gastrointestinal discomfort, social responsiveness, or anxiety, Pearson correlations were calculated to find significant trends. Microbial evenness was inversely correlated to the SCARED total score, such that lower evenness was associated with higher anxiety (Table 2). Alpha diversity was not correlated with the nutritional assessment, PGIA, SRS2, or GSRS surveys at baseline. Of the food groups assessed by the nutritional assessment survey, microbial richness was significantly positively correlated to the number of daily fruit servings (Table 2). The nutritional assessment survey was inversely correlated to the PGIA scores (Table 2), such that individuals with lower (worse) nutrition had higher (worse) ASD severity.

**Table 2.** Pearson correlations between total survey scores and alpha diversity measurements. R and p values were calculated for each of the comparisons between surveys taken by the ASD population and the microbiome diversity Shannon Index, evenness, and richness. Only significant correlations between different survey scores are listed (nutritional assessment and PGIA).

| Variable 1 | Variable 2 | R | P value |
| --- | --- | --- | --- |
| Nutritional Assessment | Shannon | 0.037 | 0.69 |
|  | Evenness | 0.015 | 0.87 |
|  | Richness | 0.13 | 0.15 |
|  | **PGIA** | **-0.19** | **0.025** |
| Nutrition - Fruit | Shannon | 0.087 | 0.35 |
|  | Evenness | 0.063 | 0.5 |
|  | **Richness** | **0.2** | **0.031** |
| PGIA | Shannon | 0.014 | 0.83 |
|  | Evenness | 0.0096 | 0.89 |
|  | Richness | 0.037 | 0.58 |
| SRS2 | Shannon | 0.046 | 0.51 |
|  | Evenness | 0.039 | 0.58 |
|  | Richness | 0.069 | 0.33 |
| GSRS | Shannon | -0.052 | 0.45 |
|  | Evenness | -0.046 | 0.5 |
|  | Richness | -0.076 | 0.27 |
| SCARED | Shannon | -0.17 | 0.051 |
|  | **Evenness** | **-0.17** | **0.045** |
|  | Richness | -0.097 | 0.26 |

### Differentially abundant microbial, pathway, and gene family abundances suggest differences in functional capacity

Microbial features represented by microbial species, pathways, and gene families significantly differed between the ASD and control cohorts. The top 50 variables of importance were determined by random forest. With additional significance testing with corrections for multiple hypotheses testing, we identified microbial features associated with the ASD or control cohorts. The mean decrease Gini values from the random forest model for the microbiome composition, pathway, and gene families are illustrated in Supplemental Figure 1. Microbes higher in abundance in the ASD cohort included *Achromobacter, Aeromonas, Bacillus sp*., *Burkholderia, Clostridium, Cronobacter, Klebsiella, Micrococcus, Mycobacterium, Rhodococcus, Shigella*, and *Streptomyces* species, some of which are pathogenic (Figure 2a). Microbes lower in abundance in the ASD cohort included *Bacillus subtilis, Faecalibacterium, Fibrobacter, Fusicatenibacter, Lachnosphira, Lactococcus, Paenibacillus, Pseudoflavonifractor, Ruminococcus*, and *Vigibacillus* species, some of which are beneficial microbes (Figure 2b).

**Figure 2.**
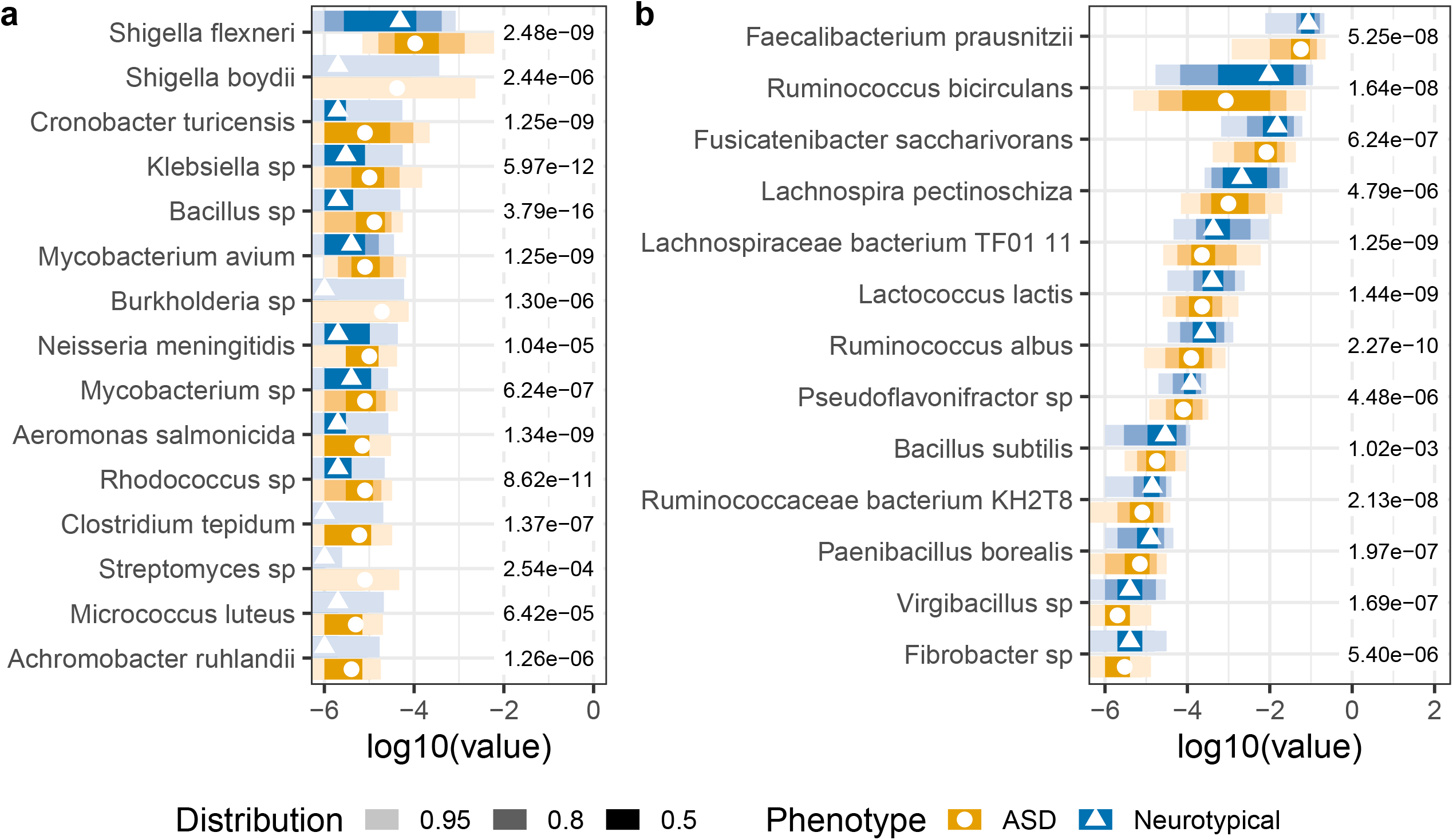
Differential abundance of microbes between ASD and NT cohorts at baseline. The top 50 microbes were selected from the variables of importance from a random forest model. Microbes listed in A) were detected at higher abundances in the ASD cohort, while microbes in B) were detected at lower abundances in the ASD cohort relative to the neurotypical cohort. Wilcoxon rank sum tests with Benjamini-Hochberg post-hoc corrections for multiple comparisons were used to determine a significant differential abundance of microbes between ASD and NT cohorts. The shading of color along the bars indicates the distribution of the data across the x-axis. Of the datapoints, 95% of the data fall within the lightest shade, 80% of the data fall within the medium shade, and 50% of the data fall within the darkest shaded bar. The different colors represent the two cohorts. The value to the right of the bars is the adjusted p-value for each test.

The MetaCyc Metabolic Pathway Database was used to annotate the functional pathway potential of the microbiome of the ASD and neurotypical cohorts. Metagenomic pathways higher in abundance in the ASD cohort were pathways involved in 1) biosynthesis of amino acids; cofactor, carrier, and vitamins; fatty acid and lipids; nucleotides; and tetrapyrrole; 2) degradation of carboxylates; and 3) fermentation of pyruvate and alcohols and fermentation to short-chain fatty acids (Figure 3a). Pathways lower in abundance in ASD were carbohydrate degradation, sulfur oxidation, and thiamine biosynthesis (Figure 3b).

**Figure 3.**
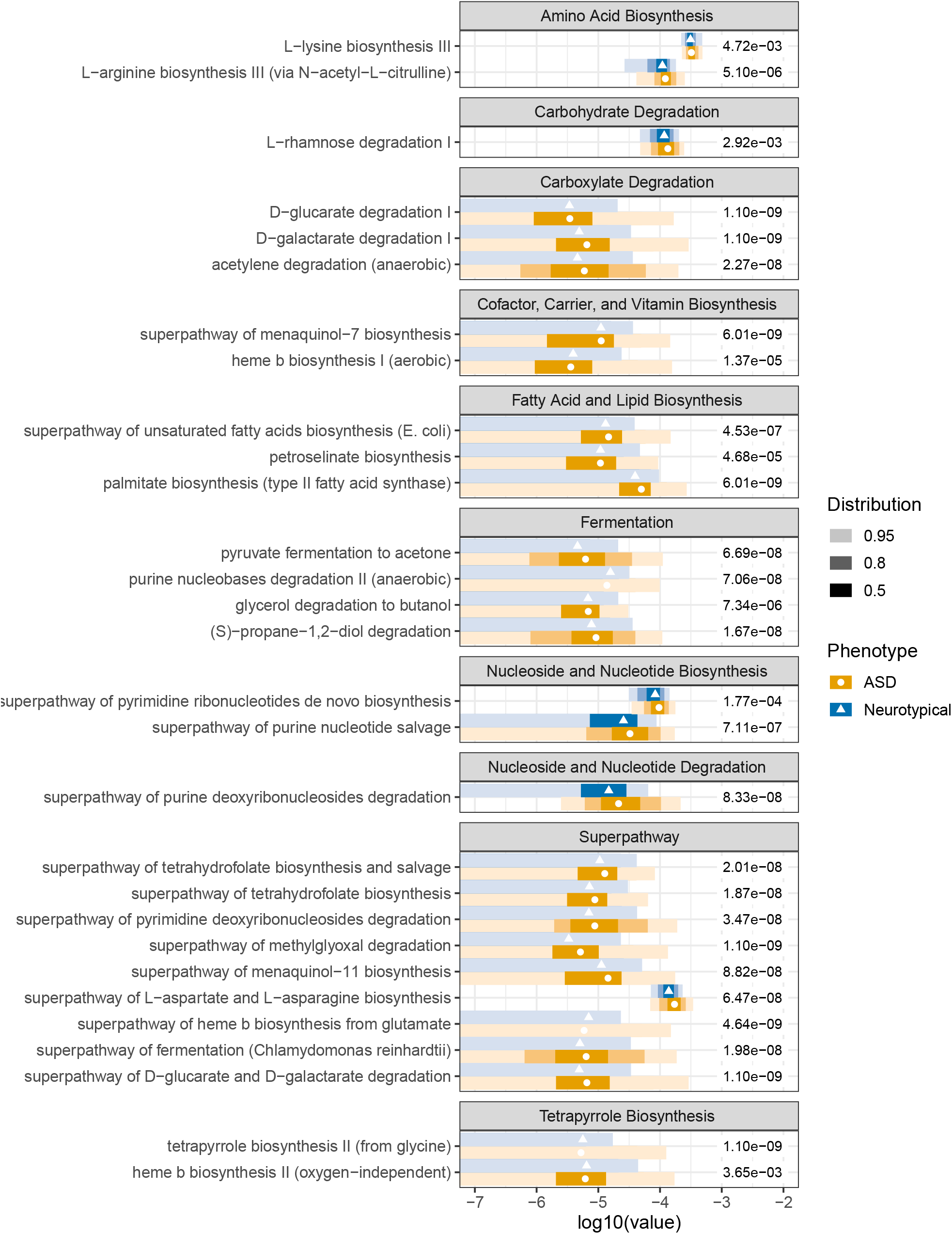
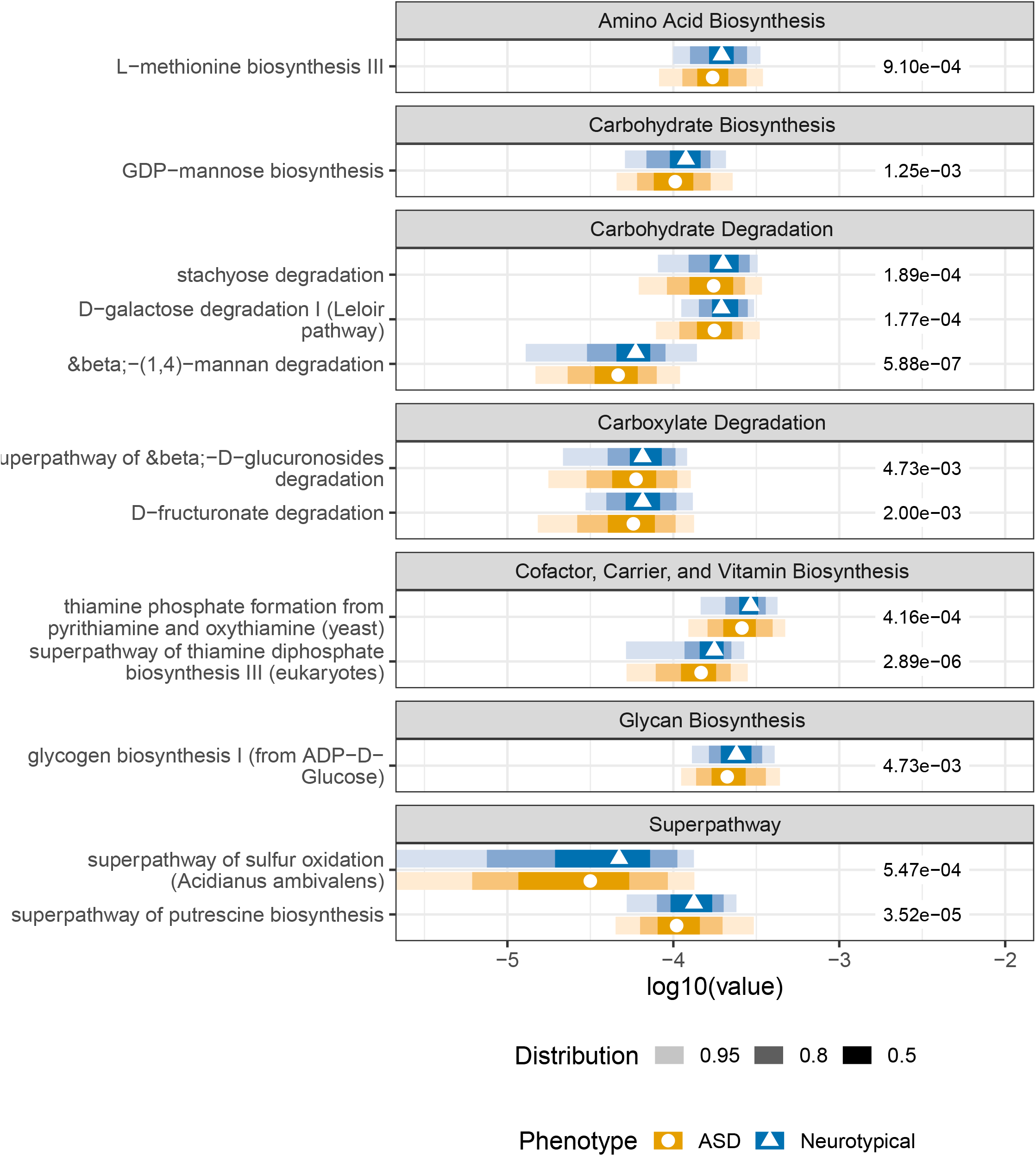
Differential abundance of pathways between ASD and NT cohorts at baseline. The top 50 pathways were selected from the variables of importance from a random forest model. Pathways listed in A) were detected at higher abundances in the ASD cohort, while pathways in B) were detected at lower abundances in the ASD cohort relative to the neurotypical cohort. Wilcoxon rank sum tests with Benjamini-Hochberg post-hoc corrections for multiple comparisons were used to determine a significant differential abundance of pathways between ASD and NT cohorts. The shading of color along the bars indicates the distribution of the data across the x-axis. Of the datapoints, 95% of the data fall within the lightest shade, 80% of the data fall within the medium shade, and 50% of the data fall within the darkest shaded bar. The different colors represent the two cohorts. The value to the right of the bars is the adjusted p-value for each test.

The UniRef (UniRef90 201901b) database was the reference database for the gene families detected in the metagenomic data. To reduce the computational burden for the statistical analyses of the gene family dataset, the sum threshold was > 1000 reads per kilobase (RPK) per gene family. Out of the top 50 gene family variables of importance, six gene families were detected with higher abundance in the ASD cohort. In comparison, 44 gene families were detected with lower abundance in the ASD cohort based on calculated mean values (Figure 4). Two annotated gene families higher in ASD were ispD and a DNA polymerase III subunit alpha (Figure 4a). Most gene families lower in ASD were annotated from *Ruminococcus spp*., *Fusicatenibacter saccharivorans*, and *Faecalibacterium prausnitzii*, which reflects the lower abundance of those microbes detected in the ASD cohort relative to the controls (Figure 2b).

**Figure 4.**
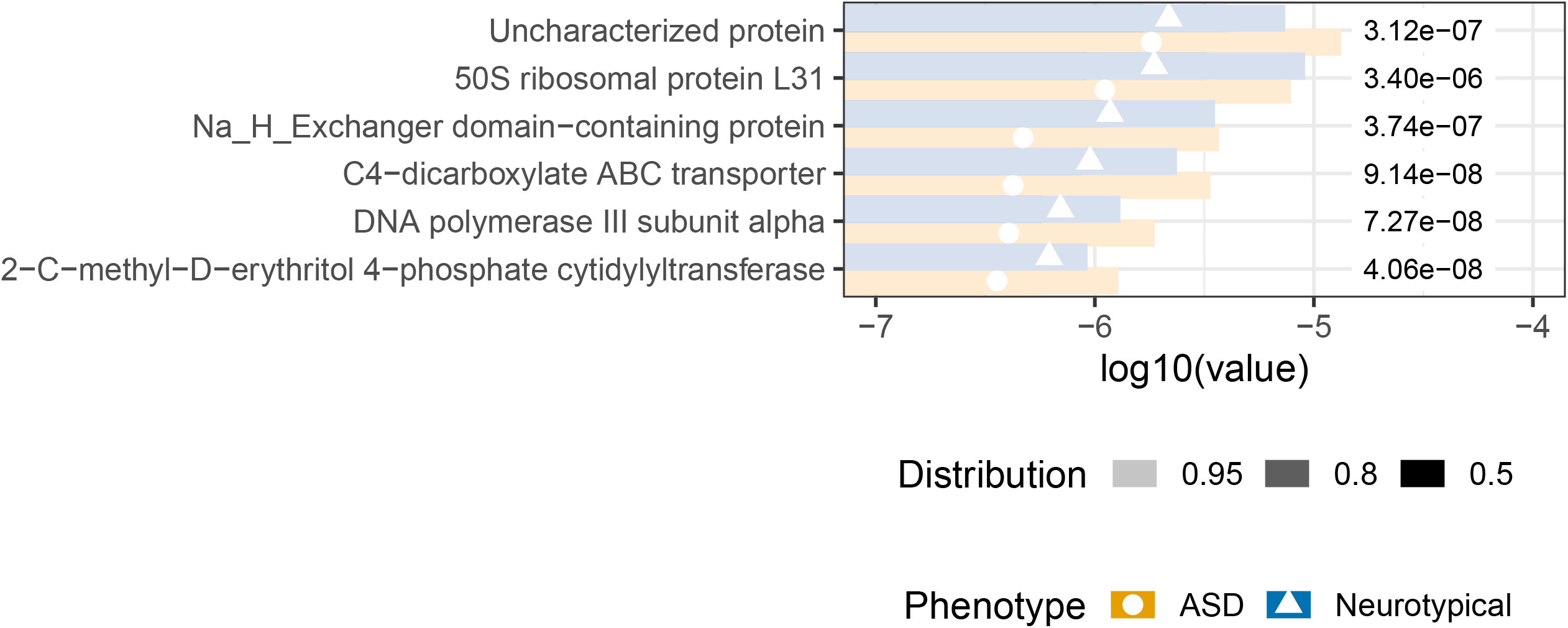
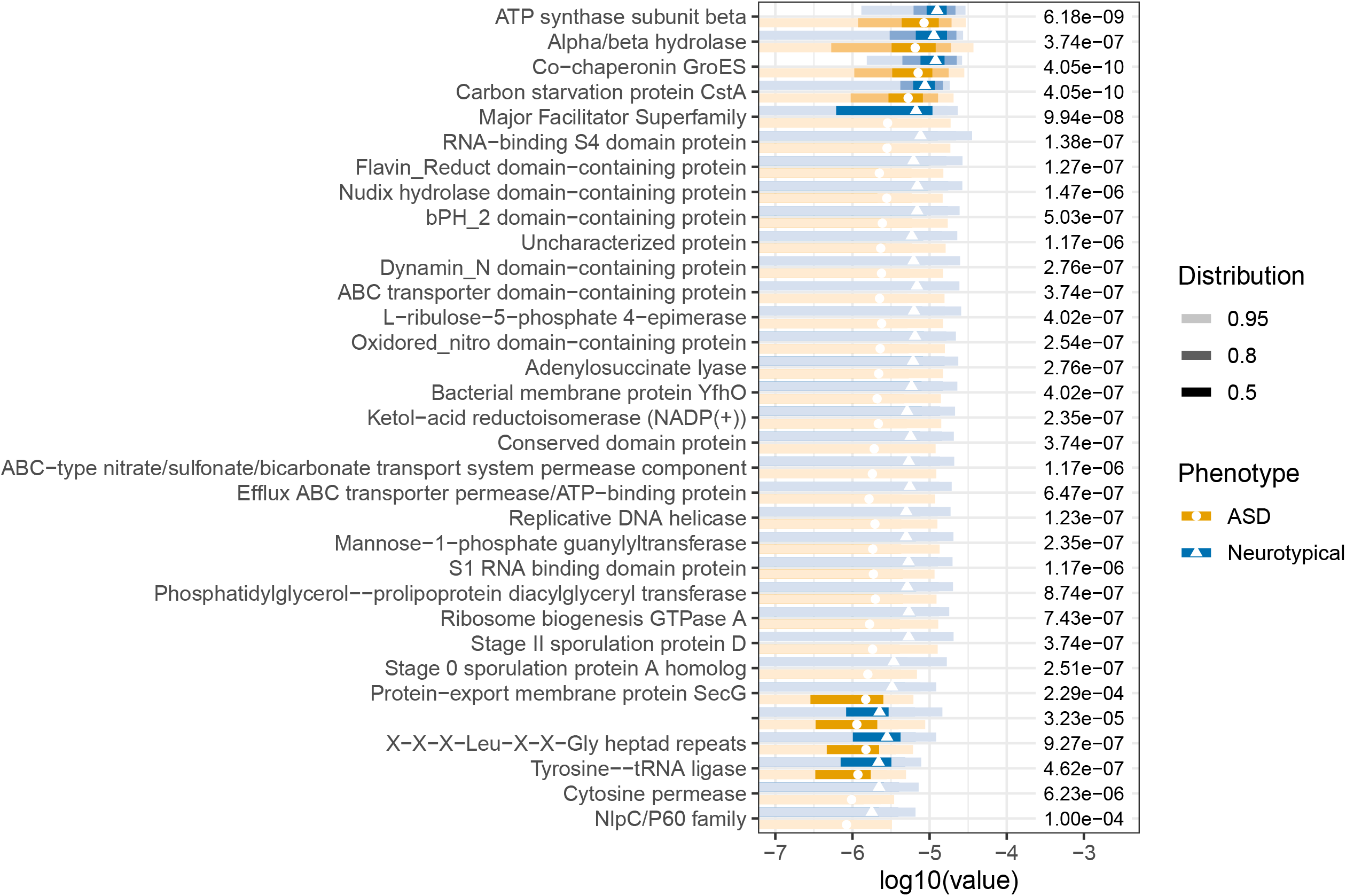
Differential abundance of gene families between ASD and NT cohorts at baseline. The top 50 gene families were selected from the variables of importance from a random forest model. Gene families listed in A) were detected at higher abundances in the ASD cohort, while gene families in B) were detected at lower abundances in the ASD cohort relative to the neurotypical cohort. Wilcoxon rank sum tests with Benjamini-Hochberg post-hoc corrections for multiple comparisons were used to determine a significant differential abundance of gene families between ASD and NT cohorts. The shading of color along the bars indicates the distribution of the data across the x-axis. Of the datapoints, 95% of the data fall within the lightest shade, 80% of the data fall within the medium shade, and 50% of the data fall within the darkest shaded bar. The different colors represent the two cohorts. The value to the right of the bars is the adjusted p-values for each test.

There were several microbes and pathways that significantly changed across timepoints in the ASD cohort (Figure 5). *Bacillus subtilis* and *Pseudoflavonifractor sp. An85* increased in abundance at timepoint 2, becoming more similar to the relative abundance of the neurotypical cohort (Figure 5a). Pathways of L-aspartate and L-asparagine biosynthesis, unsaturated fatty acids biosynthesis, purine deoxyribonucleosides degradation, and purine nucleotide salvage decreased in abundance at timepoint two relative to timepoint one and became more similar to that of the controls (Figure 5b). Petroselinate biosynthesis decreased in relative abundance between ASD timepoints 1 and 2 and was no longer significantly different than the control at timepoint 2 (Figure 5b).

**Figure 5.**
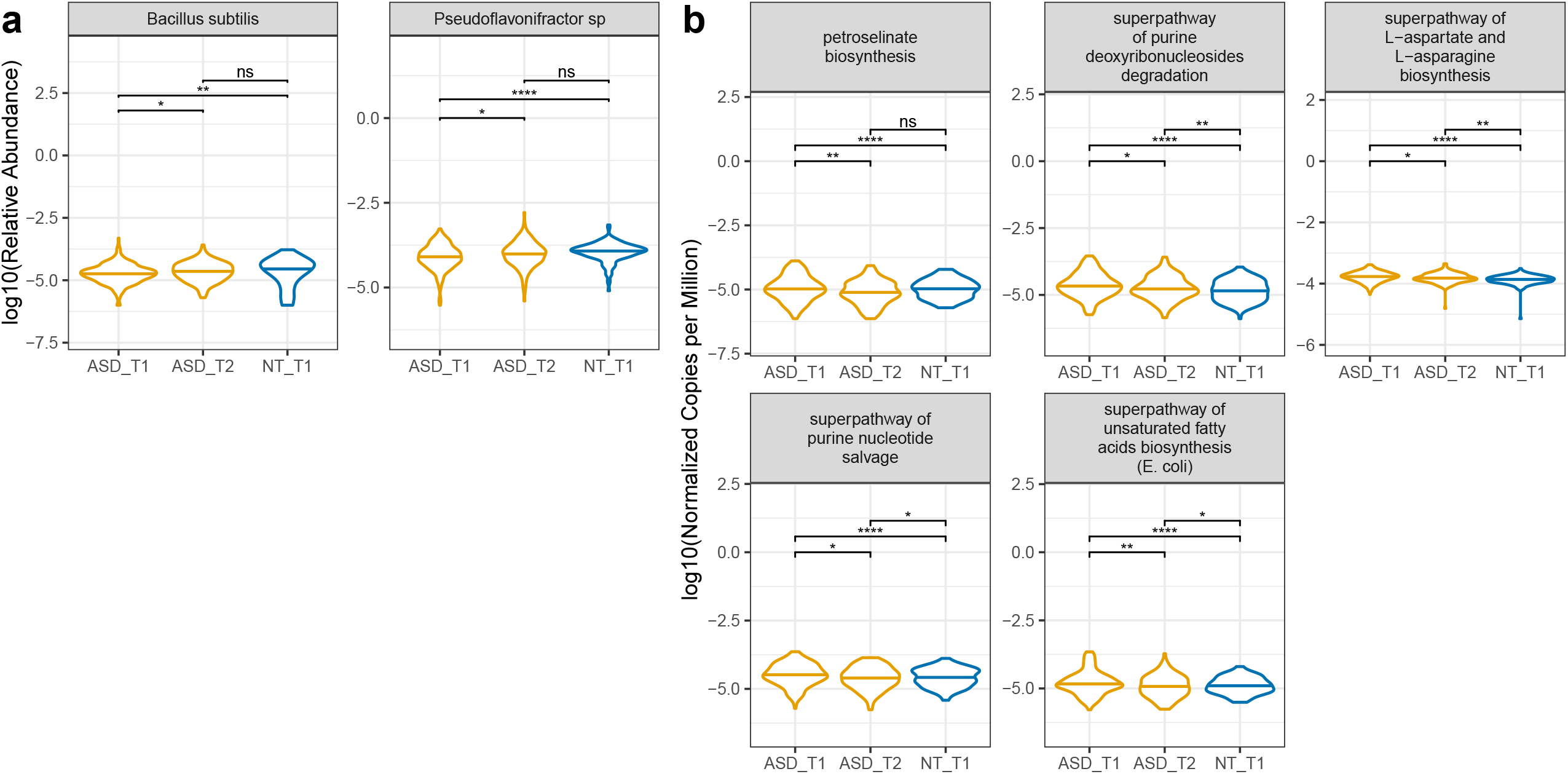
Longitudinal microbiome trends. Longitudinal assessments were made from the top 50 variables of importance obtained from the baseline comparisons of the microbiome composition, pathway abundance, and gene family datasets. ASD-paired samples, where subjects had both timepoints, were compared to the baseline neurotypical cohort. A) Microbes and B) metagenomic pathways listed were significantly different between baseline timepoint one and timepoint two in the ASD cohort. Wilcoxon tests with Benjamini-Hochberg post-hoc corrections were calculated. * p value < 0,05; ** p value < 0.01; **** p value < 0.0001; ns not significant.

### Behavioral and gastrointestinal symptom improvement with follow-up surveys

In follow-up surveys, there was reported improvement in overall autism-related symptoms and a reduction in gastrointestinal discomfort (Figure 6). For the PGIA, 62% of the subjects reported some improvement, while 30% reported no change, and 6% had slightly worsened symptoms in the question on Overall Autism and Related Symptoms (Figure 6). There was also reported improvement by at least 50% of participants in receptive language and comprehension, expressive language and speech, cognition and thinking, and gastrointestinal problems (Figure 6). The GSRS mean score significantly decreased from timepoint 1 to timepoint 2, indicating a reduction in the severity of gastrointestinal symptoms (Figure 6). There was no significant change in the SRS2 or SCARED evaluations (Figure 6).

**Figure 6.**
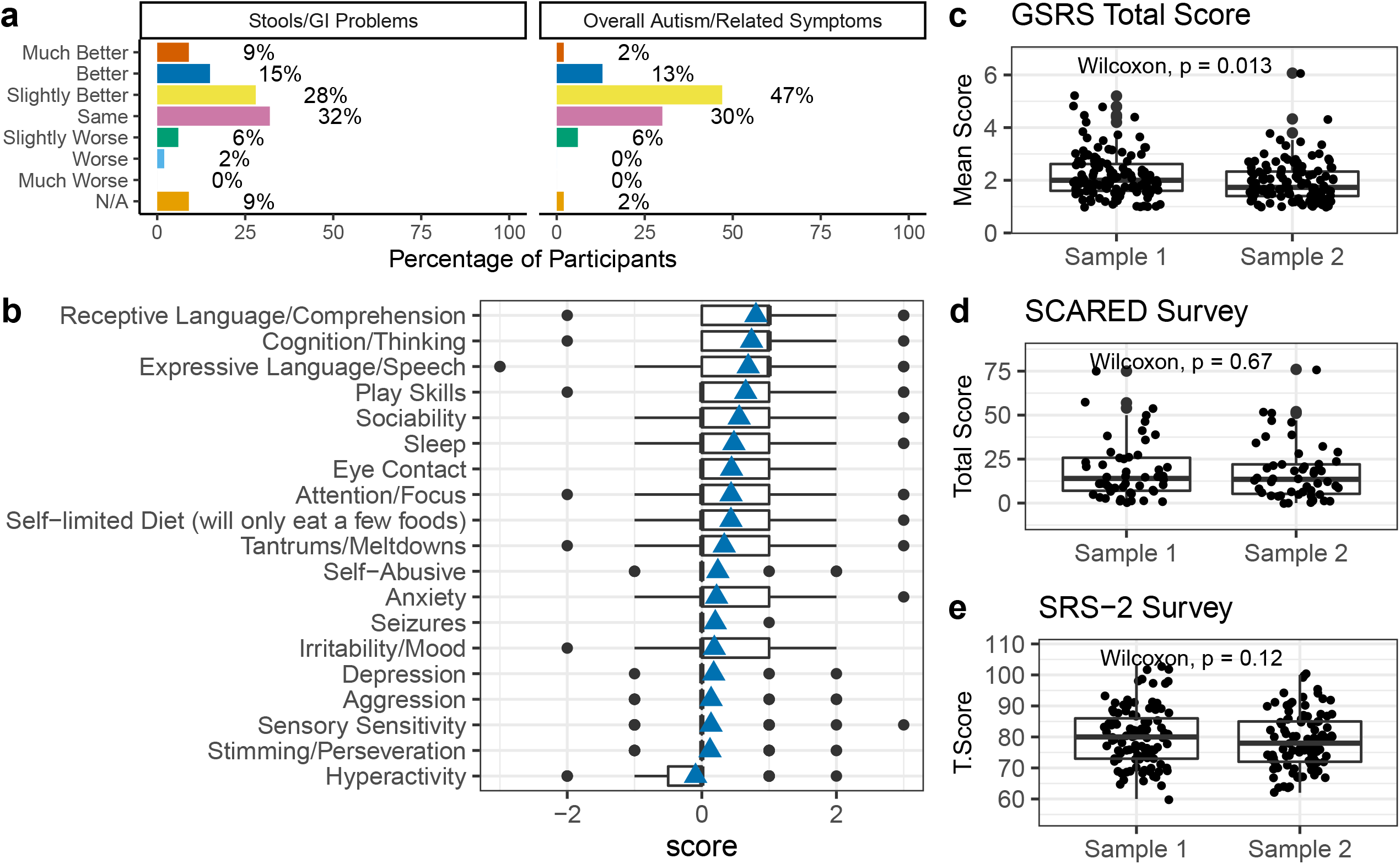
Longitudinal survey assessments. A) PGIA timepoint two survey questions regarding gastrointestinal issues and overall autism-related symptoms and the proportion of the population that saw improvement, no change, or worsening symptoms. B) The average score of the remainder of the PGIA timepoint two survey questions. The scale is −3 much worse, −2 worse, −1 slightly worse, 0 no change, +1 slightly better, +2 better, and +3 much better. The total or mean scores of the C) GSRS survey, D) SCARED survey, and E) SRS2 surveys from paired ASD samples. Paired Wilcoxon tests were performed.

## Discussion

As there has been growing evidence of the importance of the microbiome in the gut-brain axis and gut issues in ASD, identifying key microbial players may influence targets for therapeutic interventions. Although the relationship between the microbiome and ASD is multifactorial and complex, collecting behavioral, anxiety, social responsiveness, gastrointestinal, and nutritional surveys from ASD participants along with large-scale deep metagenomic sequencing may uncover signatures that may guide therapies. To our knowledge, this high-resolution method allowed for elucidating microbes, metagenomic pathways, and gene families differentially abundant between the two cohorts that are novel. Interestingly, there was no significant difference in beta diversity in any of the datasets, and most of the differences were associated with significant alpha diversity changes. In addition, there was a significant correlation or anti-correlation between species richness and daily fruit servings, the SCARED score and microbial evenness, and PGIA and nutritional assessment scores. These results indicate there are microbial features associated with ASD, in addition to the complex relationships between dietary habits, the microbiome, and ASD phenotypes.

The present finding of lower microbiome diversity in the ASD population compared to controls was consistent with some studies (D.-W. Kang et al. 2018; S. Liu et al. 2019; D.-W. Kang et al. 2013), while also contrary to some others (De Angelis et al. 2013; Zurita et al. 2020). The results of the present study indicated significantly lower levels of gut microbial alpha diversity and evenness in ASD subjects compared to neurotypical age-matched children and young adults. Still, the difference was small compared to the wide variation in alpha diversity within each group. In the De Angelis study, children were from Italy, aged 4 to 10 years old. In the 2013 Kang study, many participants had gluten and casein-free diets with nutritional supplements and probiotics (D.-W. Kang et al. 2013). In the current study, approximately a third of the participants practiced casein-free, gluten-free, or gluten and casein-free diets, while nearly 65% of the ASD participants took additional nutritional supplements. However, there was no significant difference in microbiome beta diversity based on a gluten-free diet. We also found diversity differences in pathways and gene families in ASD. Heterogeneity is also seen across geographic regions, even when controlled for DNA extraction and sequencing methods (Fouquier, Huizar, et al. 2021).

Because diet is known to contribute to microbial diversity, we investigated whether diet may be a driving force in microbial diversity in the ASD cohort. The data collected from this cohort showed no correlation between nutritional assessment and microbial diversity, similar to what was reported by (Yap et al. 2021). The nutritional evaluation showed 16.9% of the ASD cohort had excellent or very good nutrition, 21.8% had average, and 61.3% had below average or poor nutrition. Our nutritional assessment found that the majority of children with ASD had poor diets, consistent with previous findings (Zurita et al. 2020; Shmaya et al. 2015). Interestingly, we found a correlation between nutritional assessment and baseline PGIA score; but it is unclear if worse ASD symptoms result in poor diet and/or the reverse. Some habits and tendencies are characteristic of subjects with ASD, including aversions to specific types of foods and dietary intolerances (Marí-Bauset et al. 2014; Chistol et al. 2018). Sensitivities and behaviors may be correlated to nutritional choices. Chistol et al. found that autistic children with oral sensory sensitivity refused more types of foods and ate fewer vegetables.

There are similarities and differences in the gut microbiome composition of ASD found in the present study compared to the current literature. Studies have demonstrated autism gut microbiome associations with increased *Clostridiales* and *Akkermansia* and lower abundances of *Bifidobacterium, Prevotella*, and Firmicutes (Ho et al. 2020; De Angelis et al. 2013; D.-W. Kang et al. 2018; Kushak et al. 2017; L. Wang et al. 2011; H. M. Parracho et al., n.d.; Williams et al. 2011). We found higher abundances of *Shigella, Cronobacter, Klebsiella, Clostridium*, and other common pathogens and lower abundances of *Faecalibacterium, Ruminoccus, Fusicatenibacter*, and other beneficial microbes in ASD (Figure 4). These results indicate a higher abundance of pathogens and a lack of beneficial bacteria.

Microbial functional potential and metabolomics are also different between ASD and neurotypical controls. Consistent with Laue et al., we also found aspartate and asparagine biosynthesis super pathway to be higher in abundance in the ASD population (Laue et al. 2020). Metabolomics studies showed lower acetic acid and butyrate and a higher level of valeric acid in ASD (S. Liu et al. 2019). Other studies show that SCFA is higher in ASD subjects than controls, specifically acetic, butyric, isobutyric, valeric, and isovaleric acids (L. Wang et al. 2012). However, results have also been inconclusive based on direct measurements (J. Wang et al. 2019). Adams and Johansen et al. found lower levels of SCFA in ASD vs. controls (Adams, Johansen, et al. 2011). While we did not directly measure SCFA in stool, metagenomic pathways of fermentation to SCFA were higher in ASD than in controls (Figure 3b). Differences in microbial associations detected in the present study compared to previously published studies may be due to differences in size and sample population, as also seen in Fouquier et al. with study-site effects (Fouquier, Huizar, et al. 2021). Diet, therapies, autism severity, gastrointestinal discomfort, and other factors may also influence the differences in the gut microbiome seen across the ASD population.

### Stratification of the ASD population based on certain characteristics may give insight into whether there are specific microbial associations

The effect of precision synbiotic supplementation on autistic symptoms showed an overall positive response. After three months of supplementation, we obtained a second microbiome sample and collected follow-up surveys. The PGIA follow-up survey questionnaire showed an improvement in certain behaviors and the GSRS survey showed a significant decrease in GI symptom severity (Figure 6).

However, this is an open-label study, and the improvement magnitude is in the placebo effect range. We estimated the placebo effect for the PGIA by comparing against the PGIA scores of a placebo group in a 3-month randomized, double-blind, placebo-controlled trial of a vitamin/mineral supplement for children and adults with ASD (Adams, Audhya, et al. 2011). The Adams et al. study used an earlier version of the PGIA, called the PGI-R, which contained only 11 of the items on the newer PGIA. The comparison found in the Adams et al. study was that the supplement group had a PGI-R average score of 0.67 +/-0.34 and the placebo group average score of 0.34 +/-0.54 (Adams, Audhya, et al. 2011). The present study’s average PGIA score was 0.36 +/-0.55. Preclinical and clinical studies administering prebiotics and probiotics to autistic children have also demonstrated gastrointestinal and symptom improvement but have used the same single ingredient or a mix of ingredients across all participants (Santocchi et al. 2020; Y. Wang et al. 2020; Mensi et al. 2021; Eugene Arnold et al. 2019; Shaaban et al. 2018; Grossi et al. 2016b). The present results suggest that personalized synbiotics resulted in an overall change in the microbiome and its functional capacity and may contribute to improvement in autism and gastrointestinal symptoms. Although the survey to measure those improvements are comparable to a placebo effect based on the PGIA average scores, the limitations in the study, such as the lack of information on behavioral phenotypes of the control group, may also have contributed to the placebo effect. This should be controlled for in future studies. Consideration for other surveys such as CARS-2 or DSM-V may additionally clarify these results. Impactfully, 50% or more of participants reported improvement in receptive language and comprehension, expressive language and speech, cognition and thinking, and gastrointestinal problems are promising and may indicate that a synbiotic supplementation is a valuable option for children and adults with ASD. Of the 296 participants, 126 of the subjects dropped out of the study after consenting. Not all participants provided a reason for dropping out of the study, but the two primary reasons for dropping out were price (21%) and a lack of perceived benefit (34%). Three complaints were received during the study related to customer service, and the third regarded lack of perceived benefit from the supplementation.

The present study design has several strengths and limitations. Some strengths include metagenomic analysis of gut bacteria, a large sample size, an age-matched control group of healthy children and adults, two timepoints for a subset of ASD participants, and a pilot open-label treatment study. Limitations include the neurotypical control group having a different gender balance (although gender did not affect microbiome diversity) and no data on the neurotypical controls’ dietary habits, gastrointestinal symptoms, or behavior phenotypes. Without these data, we could not identify whether supplementation may or may not have altered the control cohort’s microbiome diversity, behavior, or gastrointestinal symptoms and compare the changes to the ASD cohort. However, we did uncover potential metagenomic marker genes/pathways that may influence the improvement of autistic and gastrointestinal symptoms. Overall, revealing microbiome differences in our population enables the identification of specific targets for therapeutic intervention that may aid in alleviating gastrointestinal symptoms and possibly phenotypes associated with ASD. For future studies, we plan to continue investigating the longitudinal effects of synbiotic supplementation on the microbiome and phenotypes associated with ASD.

## Data Availability

All data is proprietary to Sun Genomics.

---

**Figure S1.** Significant correlations between surveys and microbial diversity. Pearson correlations between a) the number of fruit servings and species richness, b) the SCARED total score and microbial community evenness, and c) the nutritional assessment and PGIA scores.

