## Supplementary figures and images for "Precision synbiotics increase gut microbiome diversity and improve gastrointestinal symptoms in a pilot open-label study for autism spectrum disorder"

### Supplemental Figure 1

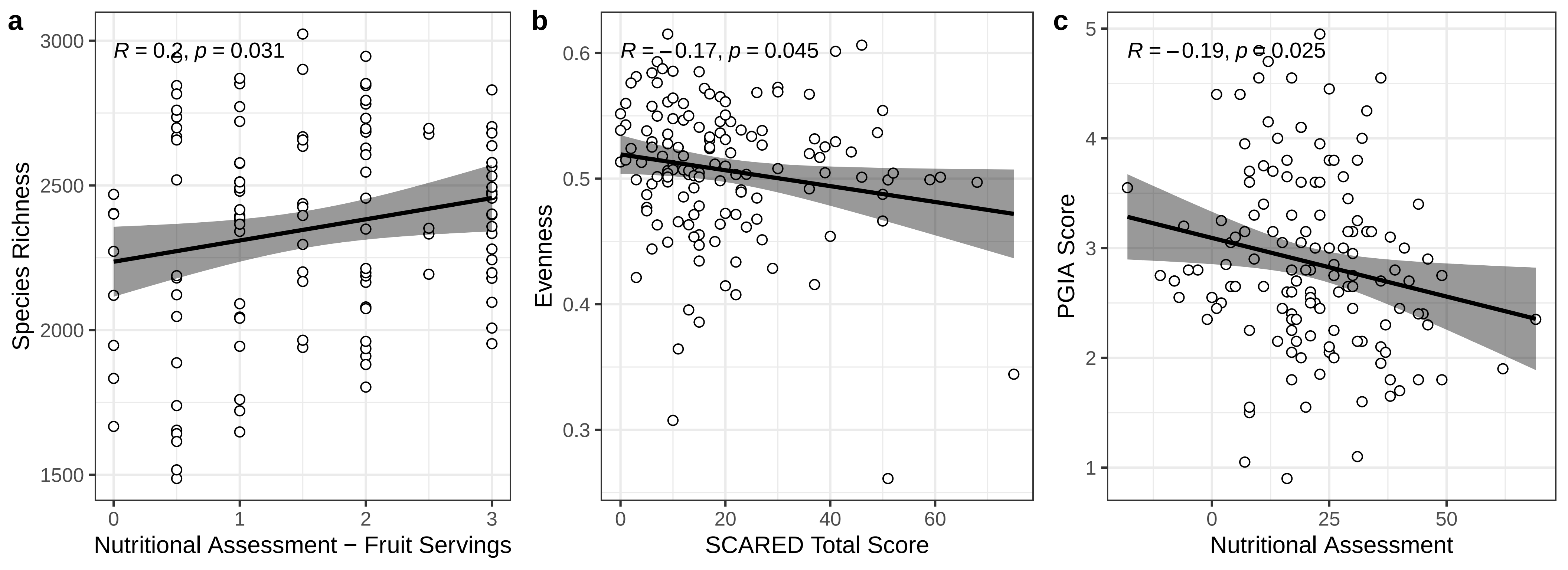
